# Evaluating any association between inflammatory diseases and connective tissue disorders with presence of atrial fibrillation/flutter

**DOI:** 10.1101/2025.02.28.25323128

**Authors:** Tala Araghi, Mehrtash Hashemzadeh, Mohammad Reza Movahed

## Abstract

**Introduction:** The role of inflammatory disease in the occurrence of atrial fibrillation/flutter (Afib/flut) is not well studied. The goal of this study was to evaluate any association between inflammatory and autoimmune disorders with the occurrences of Afib/flut using a large database

**Methods:** Using Nationwide Inpatient Sample (NIS) database and ICD-10 codes for afib/flut and inflammatory disease for the years 2016-2020, we evaluated the above association.

**Results:** A total of 23,037,013 patients were identified with a diagnosis of Afib/aflut. The following diseases were independently associated with the presence of atrial fibrillation despite adjustment for age, demographics, and traditional risk factors: Rheumatoid arthritis: OR: 1.05, CI 1.04-1.06, p<0.001, Scleroderma OR: 1.31, CI 1.26-1.36, p<0.001, Systemic Connective Tissue Disorders: OR: 1.07, CI 1.05-1.08, p<0.001, Antiphospholipid Syndrome: OR:1.36, CI: 1.31-1.42, p<001, Lupus: OR: 1.15, CI: 1.13-1.17 p<0.001 and Reynaud’s Syndrome: OR:1.1, CI: 1.07-1.13, p<0.001, Ankylosing Spondylitis was not associated with Afib/Aflut.

**Conclusion:** Using a large database, we confirmed that numerous inflammatory diseases and some connective tissue disorders are independently associated with the presence of Afib/Aflut.

## Introduction

There is a collection of heterogenous diseases referred to as immune-mediated inflammatory diseases that share a common biological mechanism related to inflammatory pathways and cytokine dysregulation^1^. Some examples of these inflammatory diseases include Rheumatoid arthritis, Scleroderma, Systemic Connective Tissue Disorders, Antiphospholipid Syndrome, and Lupus. Immune-mediated inflammatory diseases such as these often co-occur, and these patients are more likely to have another immune-mediated inflammatory disease compared to those without any^2^.

Many studies have shown a significant association between immune-mediated inflammatory diseases and rates of cardiovascular mortality for a variety of reasons such as the role of the immune system in the development of atherosclerosis^3^. Many patients with autoimmune inflammatory disease also have co-existing cardiovascular disease risk factors such as arterial hypertension and dyslipidemia^4^. Furthermore, there is also a strong association between glucocorticoid and non-steroidal anti-inflammatory drug use and cardiovascular disease; therapies such as these are often a part of management in autoimmune disease^5,6^.

Atrial fibrillation (AF) is the most common cardiac arrythmia and the incidence of Atrial fibrillation continues to increase^7^. While the incidence and prevalence of AF continue to increase, the mortality has declined^7^. Atrial fibrillation has also been found to be an independent risk factor of all-cause mortality in patients with incident AF^8^. A literature review from 2021 discussed the relationship between inflammation and the development and propagation of AF^9^. This relationship was determined when observing that inflammatory cardiac conditions such as myocarditis and pericarditis are often associated with AF^9^.

While the relationship between autoimmune inflammatory disease and atherosclerosis is well established, the relationship between autoimmune inflammatory disease and AF is not extensively studied. A recent systematic review and meta-analysis concluded that patients with inflammatory bowel disease (IBD) are at nearly 1.5 times the risk of developing AF compared to non-IBD population^10^. It is suggested that the link between IBD and AF may be related to the role of systemic inflammation^10^ and medications used to treat IBD^11^. Another systematic review concluded that patients with psoriasis are at higher risk for development of AF^12^. Our study aims to expand on this literature and evaluate any association between inflammatory and autoimmune diseases and connective tissue disorders with the presence of Atrial fibrillation and Atrial flutter using a large national inpatient database.

## Methods Data source

Patient data for this study was obtained from the Nationwide Inpatient Sample (NIS) database. Data was evaluated from the years 2016 to 2020. The NIS is a component of the Healthcare Cost and Utilization Project (HCUP), which is sponsored by the Agency for Healthcare Research and Quality (AHRQ). The NIS database accounts for admissions across approximately 1,000 hospitals. Data from the NIS database is publicly available to researchers at www.hcup-us.ahrq.gov and is de-identified. Therefore, this study was exempt from Institutional Review Board (IRB) approval.

### Data collection

The NIS database records patients’ reasons for hospitalization using ICD-10 (International Classification of Diseases, 10^th^ Revision) billing codes. In our search, we extracted relevant patient demographics by searching for specific ICD-10 billing codes. We began by identifying all patients who were admitted to NIS affiliated hospitals from 2016 – 2020. From the total patients identified, we then identified patients with a diagnosis of Atrial fibrillation and flutter and without Atrial fibrillation and flutter. The following ICD-10 codes were included in our search for AF/Flutter: I48, I48.0, I48.1, I48.11, I48.19, I48.2, I48.20, I48.21, I48.3, I48.4, I48.9, I48.91, I48.92. We also used ICD-10 codes to identify patients with specific autoimmune inflammatory diseases including Rheumatoid Arthritis (M05, M06), Scleroderma (M34), Systemic Connective Tissue Disorders (M35), Gout (M10), Psoriatic arthritis (L40.50), Polyarteritis Nodosa (M30), Reynaud’s Syndrome (I73.0), Buerger’s Disease (I73.1), Ankylosing Spondylitis (M45), Antiphospholipid Syndrome (D68.61, D68.312), and Lupus (M32.10, M32.11, M32.12, M32.14, M32.15, M32.19, M32.8, M32.9). Patients under the age of 18 were excluded from our search. A multivariate analysis was also conducted adjusting for age, gender, race, alcohol use, coronary artery disease (CAD), smoking, Type 2 Diabetes, hypertension, hyperlipidemia, Chronic Kidney Disease (CKD), and Chronic Obstructive Pulmonary Disease (COPD).

### Statistical analysis

Patient demographic, clinical, and hospital characteristics are reported as median (IQR) for continuous variables and proportions, 95% confidence intervals for categorical variables. Multivariate logistic regression was performed to ascertain the odds of binary clinical outcomes relative to patient and hospital characteristics. All statistical models will be adjusted for confounding. All analyses will be conducted following the implementation of population discharge weights. All p-values will be 2-sided and p<0.05 will be considered statistically significant. Data will be analyzed using STATA 17 (Stata Corporation, College Station, TX).

## Results

A total of 148,767,786 individuals were identified in the NIS database from the years 2016 -2020. 23,037,013 patients were identified as having a diagnosis of Atrial fibrillation or Atrial flutter. 125,730,773 patients were included who did not have AF/flutter. The mean age of patients in the study was 58.00 ± 20.23 years old. The mean age of patients with AF/flutter in this study was 74.82 ± 11.83 years old.

### Univariate Analysis

In the initial univariate analysis, several autoimmune inflammatory conditions and connective tissue disorders were associated with an increased likelihood of developing AF/flutter as indicated by the odds ratios and 95% confidence interval. AF/flutter was more prevalent in patients with Rheumatoid Arthritis, Scleroderma, Systemic Connective Tissue Disorders, Polyarteritis Nodosa, Gout, Arthropathic Psoriasis, Reynaud’s Syndrome, and Ankylosing Spondylitis. Patients with Antiphospholipid Syndrome, Lupus, and Buerger’s Disease had lower association with presence of AF/flutter.

### Multivariate Analysis

Next, a multivariate analysis was conducted adjusting for alcohol use, age, gender, race, Coronary Artery Disease (CAD), smoking, Type 2 Diabetes, Hypertension, Hyperlipidemia, Chronic Kidney Disease (CKD), and Chronic Obstructive Pulmonary Disease (COPD). The multivariate analysis showed the following odds ratios and confidence intervals: Rheumatoid Arthritis (OR: 1.05, CI 1.04 – 1.06), Scleroderma (OR: 1.31, CI 1.26 – 1.36), Systemic Connective Tissue Disorders (OR: 1.07, CI 1.05 – 1.08), Reynaud’s Syndrome (OR: 1.10, CI 1.07 – 1.13), Antiphospholipid Syndrome (OR: 1.36, CI 1.31 – 1.42), and Lupus (OR: 1.15, CI 1.13 – 1.17). Ankylosing Spondylitis was not associated with AF/flutter.

## Discussion

The relationship between autoimmune inflammatory diseases and atherosclerosis is well studied^3^, however the association between autoimmune inflammatory diseases and AF/flutter has not been well established in the literature. Atrial fibrillation is a common arrythmia that is associated with increased morbidity and mortality^13^; therefore, it is crucial to better understand who is at increased risk for developing AF/flutter. Several studies discuss the pathogenesis of AF and the role of inflammation in its pathogenesis^9,14^. Other studies have also shown that the presence of increased inflammatory markers is associated with greater AF risk^15^, further supporting the role of inflammation in the pathogenesis of AF. One proposed explanation for the role of inflammation in pathogenesis of Atrial fibrillation includes the fibrotic changes to the myocardium that result from inflammation causing the arrhythmia^9^. Given the understanding of the role of inflammation in the pathogenesis of AF it would be reasonable to suspect an association between autoimmune inflammatory diseases and AF. Our goal in this study was to show this association using a large inpatient database.

Previous literature has discussed the relationship between certain inflammatory autoimmune diseases and AF. Multiple studies have concluded that individuals with Inflammatory Bowel Disease (IBD) are at increased risk of developing AF compared to non-IBD patients^10^. Similarly, various studies have shown increased risk of new-onset AF due to psoriasis, suggesting that the proinflammatory state in psoriasis plays a role in this development^12^. The association between thyroid abnormalities and AF has also been studied, with one study suggesting that, though unclear, thyroid hormones activate various inflammatory pathways that may play a role “in the development of arrhythmogenic substrate promoting the occurrence and recurrence AF”^16^.

Additionally, a 2021 systematic review and meta-analysis of cohort studies also concluded that patients with inflammatory arthritis have increased risk of AF^17^. The paper attributes this increased risk due to the chronic inflammation reaching the same conclusions as the aforementioned studies^17^.

The result of our study adds to this growing literature showing an association between certain autoimmune inflammatory diseases and AF/flutter. Our study specifically shows an increased prevalence of AF/flutter in patients with Rheumatoid Arthritis, Scleroderma, Systemic

Connective Tissue Disorders, Reynaud’s Syndrome, and Lupus. There was no association found in this study between Ankylosing Spondylitis and AF/flutter. There are many known traditional risk factors for AF/flutter such as hypertension, CAD, and several others that we controlled for by conducting a multivariate analysis. We also controlled for certain patient demographic information including age, gender, and race within the multivariate analysis.

This study uses a large inpatient database which allows for review of a large and diverse population. However, using this database is limited by the fact that all the data comes from inpatient records and may not be reflective of the outpatient population. Additionally, with using ICD-10 codes to access our data, there is a possibility for the occurrence of inaccuracies within coding.

## Conclusion

Using a large inpatient database, we confirmed that numerous inflammatory diseases are independently associated with the presence of AF/flutter. This suggests that these inflammatory diseases are an independent risk factor for AF/flutter.

## Data Availability

NIS database is publicly available

---

**Table 1:**
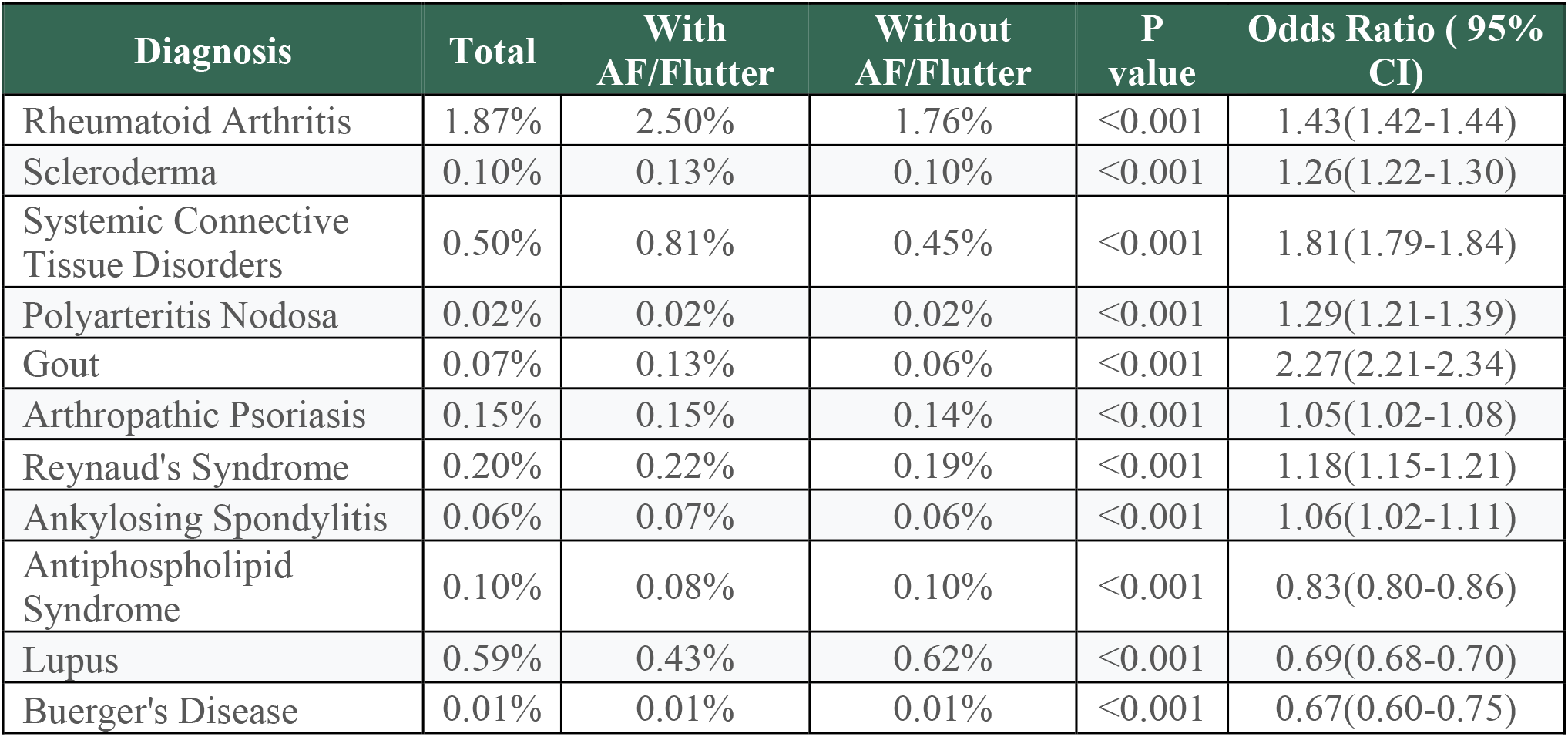
Univariate Analysis showing higher association of some inflammatory disease and connective tissue disorder with presence of atrial fibrillation/Atrial flutter except lupus, ankylosing spondylitis and Berger’s disease.

**Table 2:**
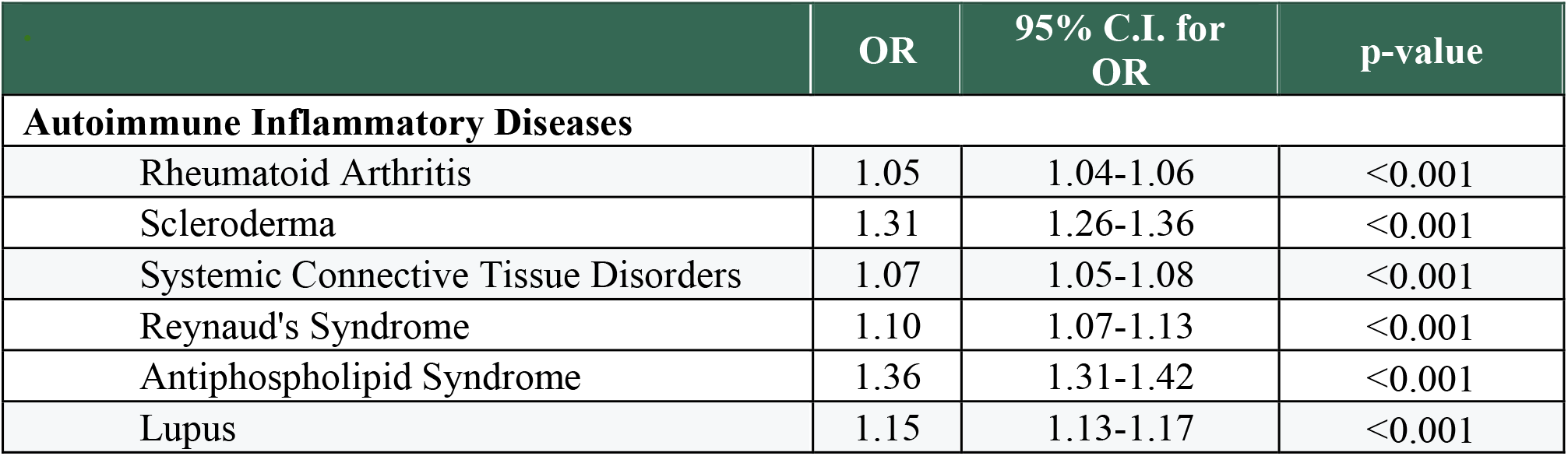
Multivariate analysis showing independent association between following inflammatory diseases and connective tissue disorder with Atrial Fibrillation/Atrial Flutter after adjusting for known risk factors, age and baseline characteristics.

|  | OR | 95% C.I. for OR | p-value |
| --- | --- | --- | --- |
| **Autoimmune Inflammatory Diseases** |  |  |  |
| Rheumatoid Arthritis | 1.05 | 1.04-1.06 | <0.001 |
| Scleroderma | 1.31 | 1.26-1.36 | <0.001 |
| Systemic Connective Tissue Disorders | 1.07 | 1.05-1.08 | <0.001 |
| Reynaud's Syndrome | 1.10 | 1.07-1.13 | <0.001 |
| Antiphospholipid Syndrome | 1.36 | 1.31-1.42 | <0.001 |
| Lupus | 1.15 | 1.13-1.17 | <0.001 |

